# Estimating population level disease prevalence using genetic risk scores

**DOI:** 10.1101/2020.02.20.20025528

**Authors:** Benjamin D. Evans, Piotr Słowiński, Andrew T. Hattersley, Samuel E. Jones, Seth Sharp, Robert A. Kimmitt, Michael N. Weedon, Richard A. Oram, Krasimira Tsaneva-Atanasova, Nicholas J. Thomas

## Abstract

Clinical classification is essential for estimating disease prevalence but is difficult, often requiring complex investigations. The widespread availability of population level genetic data makes novel genetic stratification techniques a highly attractive alternative. We propose a generalizable mathematical framework for determining disease prevalence within a cohort using genetic risk scores. We compare and evaluate methods based on the means of genetic risk scores’ distributions; the Earth Mover’s Distance between distributions; a linear combination of kernel density estimates of distributions; and an Excess method. We demonstrate the performance of genetic stratification to produce robust prevalence estimates. Specifically, we show that robust estimates of prevalence are still possible even with rarer diseases, smaller cohort sizes and less discriminative genetic risk scores, highlighting the general utility of these approaches. Genetic stratification techniques offer exciting new research tools, enabling unbiased insights into disease prevalence and clinical characteristics unhampered by clinical classification criteria.

## Introduction

The development and refinement of polygenic analysis techniques is greatly increasing our understanding of many diseases. Using polygenic risk has allowed insights into disease etiology and through Mendelian randomization evaluation of causality [1]. Clinically, capturing polygenic susceptibility through Genetic Risk Scores (GRS) can be used to determine individuals at the highest risk of a disease [2-4]. This paper concentrates on an innovative use of polygenic risk to genetically stratify a cohort into those with and without a certain disease and hence estimate disease prevalence (proportion of individuals with and without a disease). Currently estimating disease prevalence is difficult as it requires robust clinical classification. Disease specific investigations are rarely available in population level data and inaccuracies associated with self-reported diagnosis are well recognized [5, 6]. Given the increasing availability of population level genetic data, novel polygenic estimates of disease prevalence are an extremely attractive alternative.

The basis of genetically determining disease prevalence is fundamentally that the distribution of a specific disease GRS within a population will reflect the mixture of genetic risk scores of those with the disease (cases) and those without (non-cases). This mixture GRS distribution will lie between reference populations of cases and non-cases and will reflect the relative proportion of cases to non-cases (Fig. 1). The location of the mixture population’s GRS distribution in comparison to the GRS distribution of known cases and non-cases allows the respective proportion of each group to be determined which provides a genetic-based estimate of disease prevalence. Furthermore, using the genetically calculated proportion of a disease within a population allows the additional benefit of associated clinical features of the genetically defined disease cohort to be determined. It is worth emphasising that in almost all polygenic risk situations, even those at the highest genetic risk are unlikely to develop the relevant disease and therefore this concept does not remain valid at an individual level. Nonetheless, at a population level the average genetic risk score will be higher in a cohort with disease versus those without.

**Figure 1:**
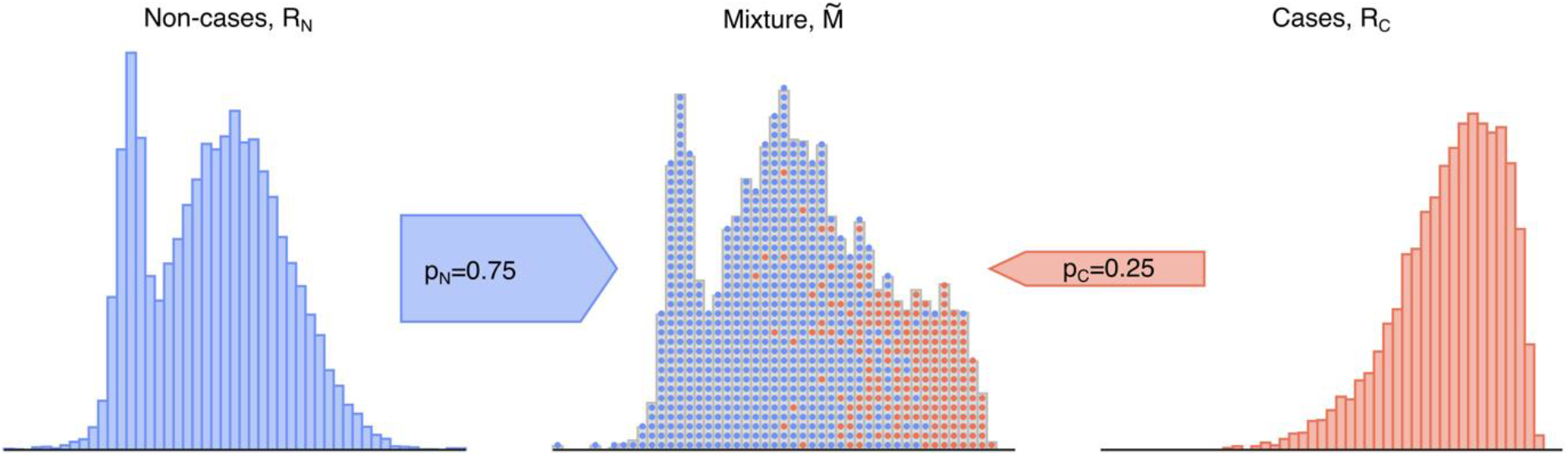
Illustration of a mixture population drawn from two reference populations. This emulates the real-world scenario of a population composed solely of individuals drawn from each subpopulation of non-cases (*R*_*N*_, blue) and cases (*R*_*C*_, red). Data are sampled with replacement from the two depicted reference cohorts to generate a mixture 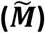 cohort of a particular sample size and proportion, which possesses features of both reference cohorts. Figure generated using artificial data.

**Figure 2:**
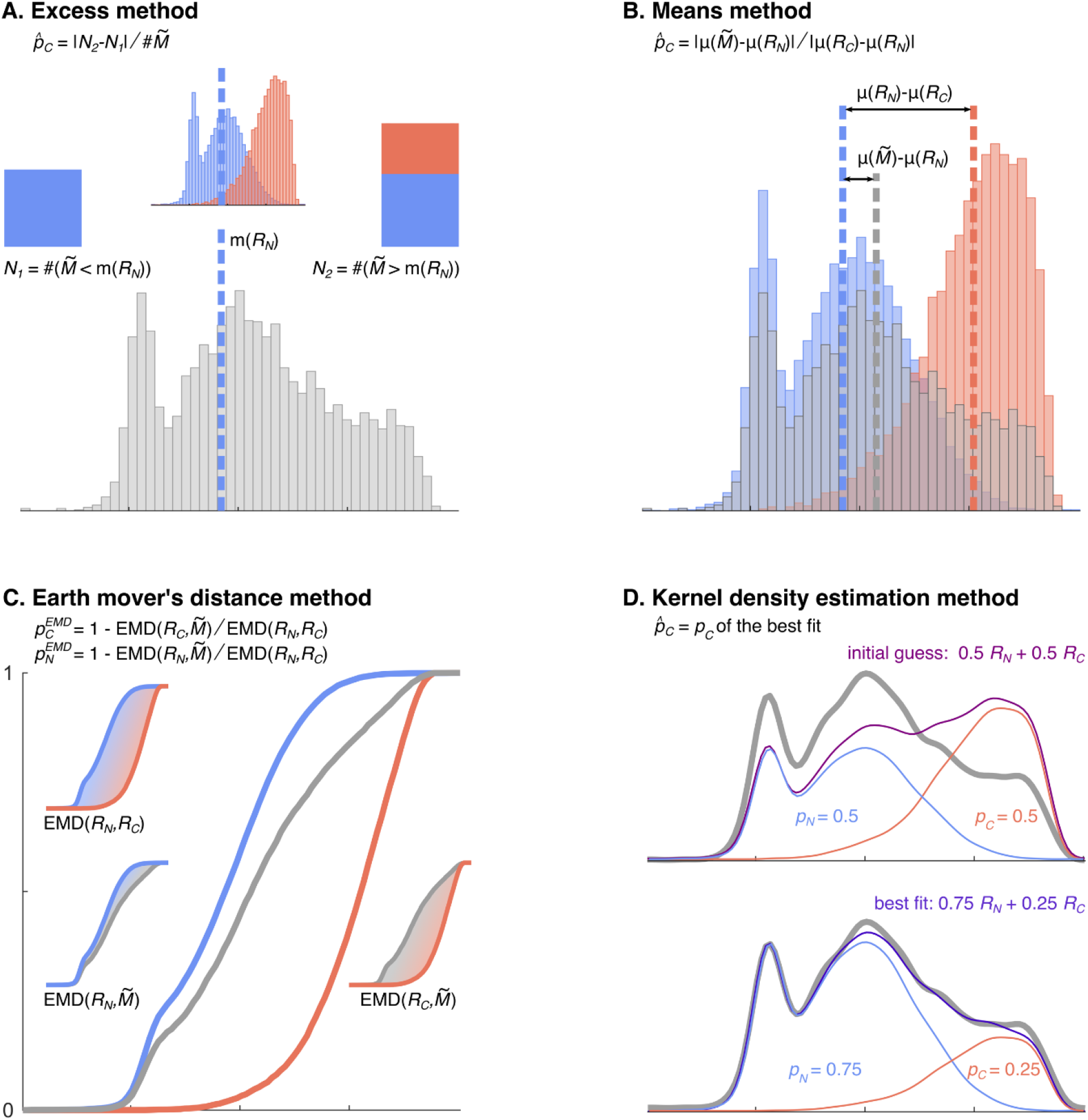
Illustration of the four proportion estimation methods. Each method uses different characteristics of the mixture and reference cohorts to estimate the proportion of constituents of the mixture cohort (*p*_C_ and *p*_N_). The Excess method (A) considers the number of cases above the mixture median in excess of those expected in a pure control (non-cases) reference cohort. The Means method (B) uses the normalised difference of the mixture cohort’s mean and the two reference cohorts’ means. The Earth Mover’s Distance method (C) uses the weighted costs of transforming the mixture distribution into the reference distributions. The Kernel Density Estimation method (D) fits smoothed templates to each of the reference distributions and then fits a weighted sum of these templates to the mixture distribution, adjusting the amplitudes of each with the Levenberg–Marquardt algorithm. Figure generated using artificial data.

In this paper we assess, the performance and utility of polygenic stratification as a tool for determining disease prevalence. Through simulated scenarios and real-world data, we evaluate different mathematical techniques for determining disease prevalence based on the GRS distribution within a cohort. The generalizability and robustness of genetic stratification has been investigated through a systematic evaluation of the cohort characteristics required for estimates to remain robust. Specifically, the impact on performance of the prevalence of the disease, the mixture cohort size and the strength of genetic predisposition for a disease. Finally, in order to highlight the utility of the proposed framework we apply our methodologies in the context of identifying the prevalence of undiagnosed coeliac disease within a cohort adhering to a gluten-free diet.

### Genetic stratification summary

We present three methods developed to estimate the proportions of cases and non-cases in an unknown mixture cohort using GRS distributions and compare them with the published Excess approach [7]. The methods’ performance characteristics are evaluated over clinically relevant parameter ranges using genetic risk scores for type 1 diabetes (T1D), type 2 diabetes (T2D) and coeliac disease, as well as synthetic data. Clinical sample sets were taken from the following cohorts: T1D (n=1,963) and T2D (n=1,924) from the Wellcome Trust Case Control Consortium (WTCCCC) [8], Coeliac disease reference cases (n=12,018) from a combination of European studies [15] with non-cases (controls) and mixture (gluten-free diet) cohorts from UK Biobank (n=12,000 and n=12,757, respectively) [9].

To compare the methods under different conditions, the T1DGRS data were split in half to form reference cohorts and an independent hold-out set for generating parameterised mixture cohorts (Figs 3 and 4). In these analyses, mixtures were constructed by sampling with replacement, enabling larger mixture sizes to be used than the size of the hold-out sets from which they were derived. Synthetic data sets were also constructed from Gaussian distributions of equal standard deviations (set to 1) but different means (see Table 1 and Fig 5). For the reference cohort of cases, *R*_*C*_, the mean of the generating distribution was always 0 while the mean for the non-cases cohort, *R*_*N*_, was systematically varied in order to investigate the effect of differences in discriminability signified by the area under the curve (AUC) of the GRS distribution. For further details of the data sets, see Materials and Methods.

**Figure 3:**
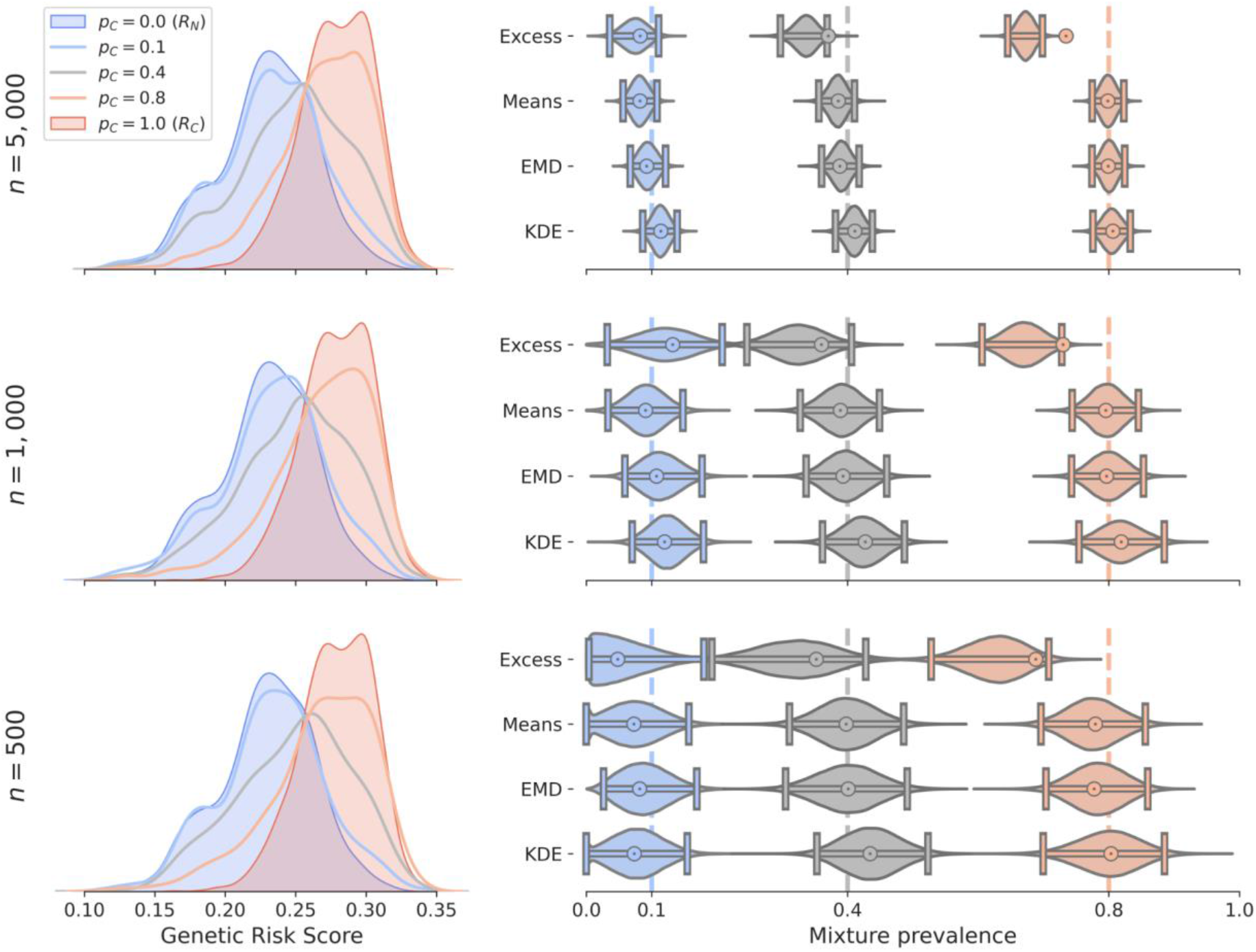
A comparison of the four methods prevalence estimates and confidence intervals for varying proportion of cases and for three sample sizes. Mixture distributions of non-cases and T1D patients from WTCCC [8] were constructed with *p*_*C*_ = {0. 1, 0. 4, 0. 8} (shown in blue, grey and red respectively) and *n* = {500, 1000, 5000}. (Left column) The constructed mixture distributions and reference distributions (*R*_*C*_, shaded red and *R*_*N*_, shaded blue) from which they were constructed. (Right column) Prevalence estimates, 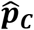 (bullseye) obtained by each of the four methods for varying *p*_*C*_ (x-axis) and cohort size, *n* (rows). Each estimated 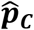 value is shown together with a violin plot illustrating the distribution of the 100,000 estimates of prevalence 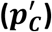 in the bootstrap samples and with confidence intervals (*α* = 0. 05) shown as horizontal lines with vertical bars at the ends. Dashed vertical lines indicate reference prevalence values *p*_*C*_. In all the cases, for the Excess method we observe a large offset between the violin plots (including confidence intervals) and the 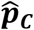 value. This offset is a result of the systematic bias of the Excess method. The other three methods generally show much less bias. Sample sizes: *R*_*C*_ – cases WTCCC T1D (*n* = 982), *R*_*N*_ – non-cases WTCCC T2D (*n* = 962), mixtures – sampled with replacement from a holdout half of the *R*_*C*_ (*n* = 981) and *R*_*N*_ (*n* = 962) samples.

**Figure 4:**
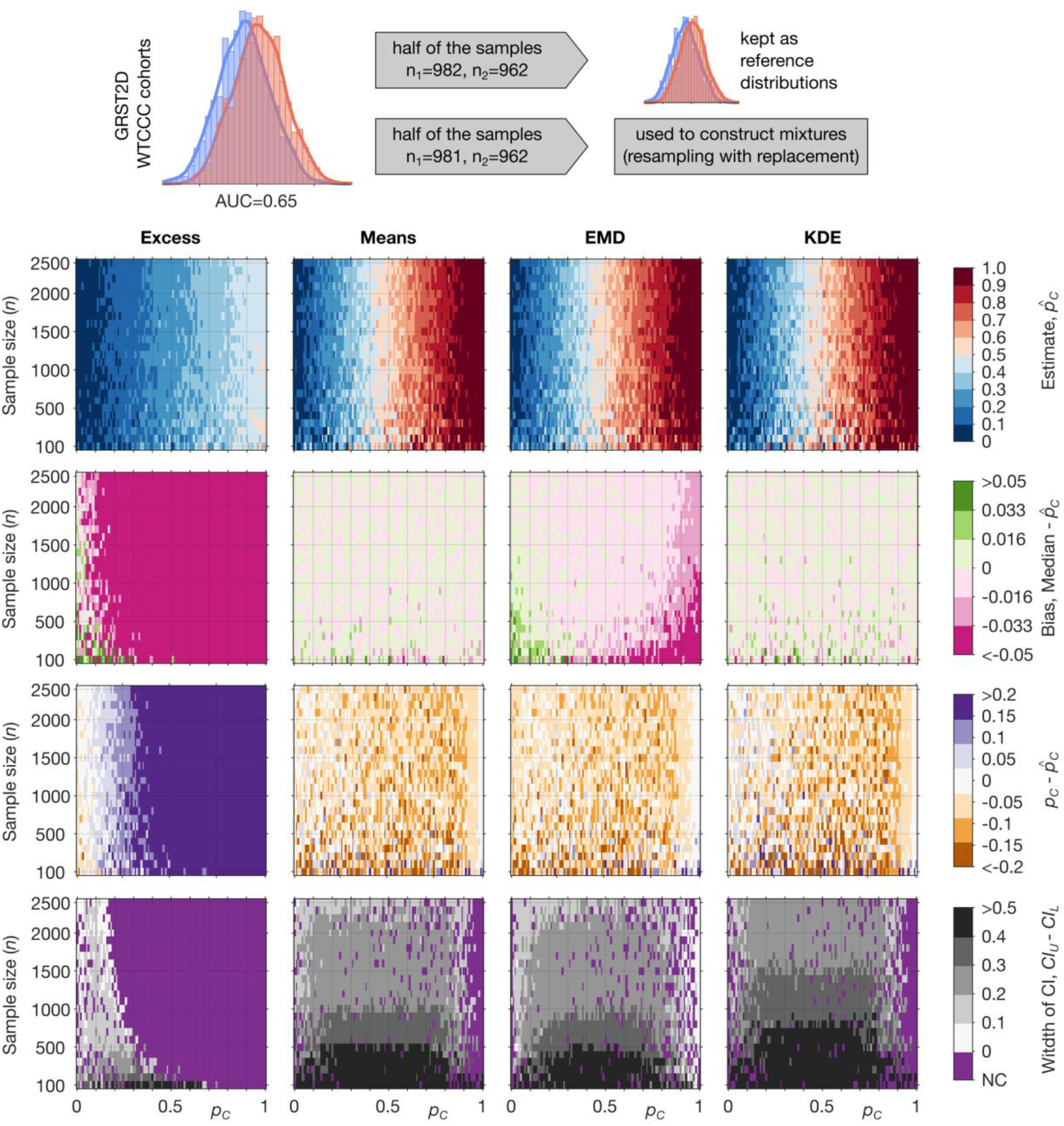
A comparison of the four methods with prevalence estimates and confidence intervals for varying proportion of disease and cohort sizes using the (Type 2 GRS) from the WTCCC dataset: T1D (n = 1, 963), T2D (n = 1, 924). (Top row) Estimate of prevalence 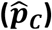 in the constructed mixtures. (Second row) Bias of the prevalence estimates 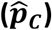 across the constructed mixtures. (Third row) Deviation from the true proportion 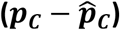 across the constructed mixtures. (Bottom row) The width of confidence intervals (*CI*_*U*_ − *CI*_*L*_) of the estimates across the constructed mixtures. The purple colour (bottom row) indicates regions in which the confidence interval did not include the true value (*p*_*C*_), *CI*_*L*_ = *CI*_*U*_ or the confidence interval was undefined (both latter cases can happen if 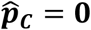 or 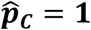). Sample sizes: *R*_*C*_ – cases WTCCC T1D (*n* = 982), *R*_*N*_ – non-cases WTCCC T2D (*n* = 962), mixtures – sampled with replacement from a hold-out half of the *R*_*C*_ (*n* = 981) and *R*_*N*_ (*n* = 962) samples; see Materials and Methods for details.

**Table 1.** The minimum mixture size required to give precision of +/- 0.05 around a prevalence of 0.1 with increasing AUC. The table shows the median minimum mixture size, 25% quantile, 75% quantile, and the number of misses (coverage probability) – when the confidence interval at the minimum mixture size did not contain the true prevalence value (*p*_*C*_ = 0. 1); the minimum mixture size is based on 100 estimation runs (see Materials and Methods – Varying mixture size). The estimates based on the Excess method do not converge to *p*_*C*_ = 0. 1 with increasing sample size for AUC = {0.6, 0.65, 0.7, 0.75}. The number of misses quickly increases to 100, showing that the estimates converge to a value much smaller than 0.1. The estimates based on the KDE method converge to *p*_*C*_ = 0. 1 for AUC = 0.6. For further details of the computations see Materials and Methods.

| Method/AUC | 0.6 | 0.65 | 0.7 | 0.75 | 0.8 | 0.85 | 0.9 |
| --- | --- | --- | --- | --- | --- | --- | --- |
| **Excess** | - | - | - | - | 3200 | 3100 | 3100 |
|  | Q25: - | Q25: - | Q25: - | Q25: - | Q25:3000 | Q25:2900 | Q25:3000 |
|  | Q75: - | Q75: - | Q75: - | Q75: - | Q75:3400 | Q75:3300 | Q75:3200 |
|  | 87/100 | 62/100 | 42/100 | 19/100 | 13/100 | 2/100 | 3/100 |
| **Means** | 26500 | 11500 | 6200 | 3700 | 2500 | 1700 | 1100 |
|  | Q25:25850 | Q25:11100 | Q25:6000 | Q25:3600 | Q25:2400 | Q25:1600 | Q25:1100 |
|  | Q75:27300 | Q75:11700 | Q75:6375 | Q75:3900 | Q75:2525 | Q75:1700 | Q75:1200 |
|  | 4/100 | 1/100 | 1/100 | 0/100 | 3/100 | 4/100 | 1/100 |
| **EMD** | 25500 | 10800 | 5700 | 3400 | 2200 | 1500 | 1000 |
|  | Q25:24600 | Q25:10400 | Q25:5550 | Q25:3300 | Q25:2100 | Q25:1400 | Q25:1000 |
|  | Q75:26300 | Q75:11200 | Q75:6000 | Q75:3600 | Q75:2300 | Q75:1500 | Q75:1000 |
|  | 0/100 | 0/100 | 0/100 | 1/100 | 0/100 | 2/100 | 0/100 |
| **KDE** | 38250 | 17000 | 9000 | 5500 | 3400 | 2100 | 1300 |
|  | Q25:37000 | Q25:16000 | Q25:8500 | Q25:5300 | Q25:3300 | Q25:2100 | Q25:1300 |
|  | Q75:39500 | Q75:18000 | Q75:9125 | Q75:5700 | Q75:3500 | Q75:2200 | Q75:1400 |
|  | 54/100 | 17/100 | 15/100 | 4/100 | 3/100 | 0/100 | 0/100 |

**Figure 5:**
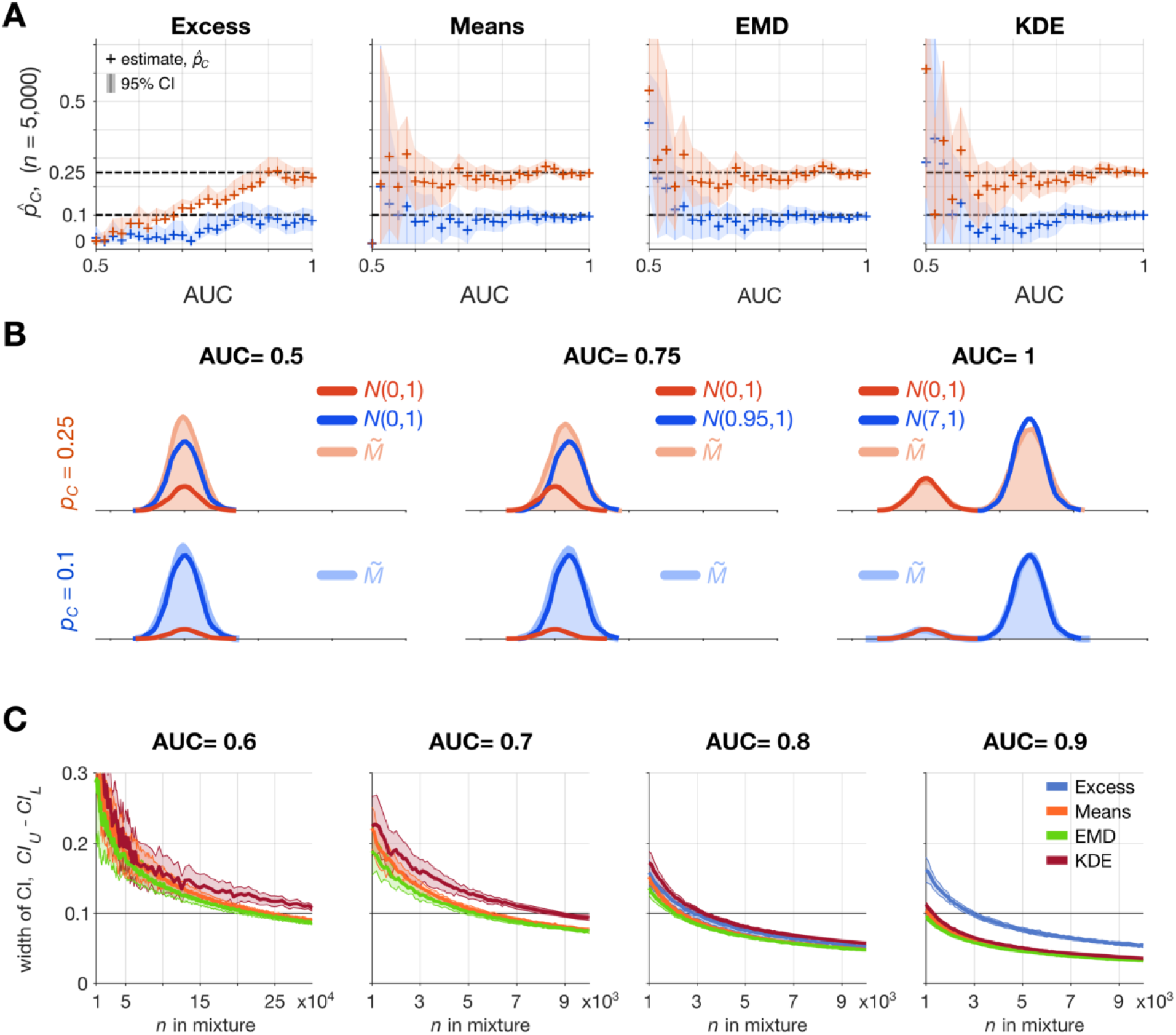
A comparison of the four methods using an artificial genetic risk score with increasing discriminative ability as measured by AUC, from AUC = 0.5 (no discriminative ability) through to AUC = 1, (complete differentiation). (A) The estimated proportion with confidence intervals around *p*_*C*_ = 0. 1 (blue) or *p*_*C*_ = 0. 25 (red) for each of the methods (Excess, Means, EMD, KDE) are shown using mixture size, *n* = 5000. (B) The constructed mixture distributions and reference distributions (*R*_*C*_, shaded red and *R*_*N*_, shaded blue) from which they were constructed for AUC = {0.5, 0.75, 1}. (C) Dependence of the width of CI (*CI*_*U*_ − *CI*_*L*_) on the number of points in the mixture sample for AUC = {0.6, 0.7, 0.8, 0.9} and *p*_*C*_ = 0. 1. Curves and shading show median +/- standard deviation of the width of CI. The plot for the Excess method for AUC = {0.6, 0.7} is omitted because the method does not converge to *p*_*C*_ = 0. 1. This figure is generated using artificial data: *N*(μ,σ) is a normal distribution with mean μ = {0.0, 0.08, 0.15, 0.22, 0.29, 0.37, 0.44, 0.51, 0.59, 0.66, 0.74, 0.82, 0.91, 0.99, 1.09, 1.19, 1.29, 1.4, 1.52, 1.65, 1.81. 1.98, 2.19, 2.47, 2.91, 7} and standard deviation σ = 1 and 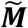 is a mixture of the two normal distributions (*R*_*C*_ is always *N*(0,1)). Both reference samples have *n* = 2000. For AUC=0.5, means of the constructed mixture samples (for *p*_*C*_ = 0. 1 and *p*_*C*_ = 0. 25) were smaller than both means of the reference samples, in these cases the prevalence estimate from the Means method is assumed to be 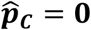 and confidence intervals are undefined due to undetermined acceleration value.

In each method, two cohorts consisting of the genetic risk scores of individuals with and without a particular polygenic disease were taken as references, denoted *R*_*C*_ (the reference cohort of cases) and *R*_*N*_ (the reference cohort of non-cases). The proportions of individuals from these reference cohorts (denoted *p*_*C*_ and *p*_*N*_ respectively) who comprise an unknown mixture cohort 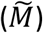 were estimated based on the properties of the reference cohorts. When only one proportion is mentioned, this is *p*_*C*_ (i.e. relative to the reference cohort of cases, *R*_*C*_), unless otherwise stated. The cohort characteristics used are dependent upon the particular method employed as illustrated in Fig. 2 and are detailed below^1^.

Throughout this paper we assume that the unknown mixture cohort is composed solely of the samples that come from the two reference cohorts (blue and red dots in Fig. 1). In practice, this means that *p*_*N*_ (prevalence of non-cases) and *p*_*C*_ (prevalence of cases) sum to one, *p*_*N*_ + *p*_*C*_ = 1, and so accordingly, the proportion of non-cases was calculated as: *p*_*N*_ = 1 − *p*_*C*_.

Furthermore, the presented Earth Mover’s Distance (EMD) and Kernel Density Estimation (KDE) methods make it possible to check if this assumption is satisfied. We revisit details of such checks in the discussion and supplementary information.

Finally our methods are all based on the assumption that cases and non-cases are genetically equivalent between the reference and mixture cohorts. This assumption must hold true for estimates to be valid. This is of particular importance when studying different geographical populations where allele frequencies are known to vary [10, 11]. To ensure this assumption is fulfilled the selection criteria and demographics of the reference and mixture cohorts should be compared to ensure equivalence. In this manuscript all analyses are restricted to white Europeans, reflecting the populations that the reference GRS distributions were derived from. Furthermore where possible the GRS of the reference non-cases (controls) and cases should be compared with the GRS of non-cases and cases within the same population the mixture has been taken from. This could be done, for example, by means of a statistical test appropriate for assessment of the observed GRS distributions. An example of the importance of this is the T2DGRS for a reference T2D population from the WTCCC [8]. The WTCCC cohort was largely selected based on a positive family history of T2D or early disease onset and is therefore enriched for T2D risk variants. As shown in Supplementary Fig. 1 the distribution of T2DGRS of unselected T2D cases from population data in UK Biobank is significantly lower than the T2DGRS in the WTCCC T2D reference. The T2DGRS in UK Biobank population T2D cases only becomes equivalent to the WTCCC when the same case selection criteria are mirrored. If this WTCCC cohort was used as a reference T2D population when evaluating prevalence of T2D in UK biobank, it would have influenced the accuracy of estimates since it is not reflective of an average T2D cohort.

### The Excess method

This estimates the proportion from the number of excess disease cases above the mixture cohort’s median score compared to the equal numbers expected in a pure control cohort (Fig. 2A). We illustrate the method as introduced in [7].

### The Means method

This compares the mean genetic risk score of the mixture cohort to the means of the two reference cohorts and estimates the mixture proportion according to the normalised difference between the two (Fig. 2B).

### The Earth Mover’s Distance (EMD) method

This uses the weighted cost of transforming the mixture distribution into each reference distribution (more formally, the integral of the difference between the cumulative density functions, i.e. the area between the curves). This method allows *p*_*N*_ and *p*_*C*_ to be computed independently (Fig. 2C) and so provides a way to validate the assumption that the mixture is composed solely of the samples from the two reference cohorts, 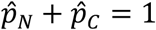; if the sum is significantly different from 1, then the assumption is not satisfied. In this study we use the mean of the two estimates for 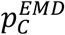 and 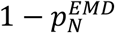 as the estimate of the 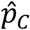.

### The Kernel Density Estimation (KDE) method

This method fits a smoothed template to each reference distribution (by convolving each sample with a Gaussian kernel) and builds a model of the mixture as a weighted sum of these two templates. The method then adjusts the proportion of these templates with the Levenberg–Marquardt (damped least squares) algorithm until the sum optimally fits the mixture distribution (Fig. 2D), noting that the algorithm could find one of potentially several local minima. In other words, the method finds (one of) the linear (convex) combination(s) of the reference distributions that best fits the mixture distribution.

## Results

### Performance of genetic stratification

We start by using the T1D genetic risk scores (AUC = 0.88) to evaluate the performance of all four methods on artificially constructed (synthetic) mixtures. The mixtures are generated by sampling with replacement from half of the reference data (holdout subset), to ensure the reference and mixture cohorts are independent and identically distributed (for details see Materials and Methods). Figure 3 demonstrates that genetic stratification allows robust estimates of disease prevalence (proportion of cases to non-cases) around known values. The accuracy (defined as deviation from the true proportion) and precision (defined as confidence interval width) of estimates is dependent on the following variables: proportion of cases and non-cases within the mixture, the mixture size and the discriminative ability of the GRS. For each method we describe how each of these variables affects the accuracy and/or precision of prevalence estimates.

### What is impact of the proportion of cases to controls in the mixture cohort?

In all methods except the Excess, away from extremes of proportion, varying the proportion of cases to controls has no impact on the accuracy or precision of prevalence estimates (Fig. 3). Using heat maps we illustrate the combined effect of gradually changing both proportion and mixture size on the accuracy of estimates (Fig. 4 and Supplementary Fig. 2). At extremes of proportion accuracy significantly reduces, tending to underestimate at high proportions and overestimate at low proportions. Increasing sample size reduces the extent to which proportions are classed as extremes thereby improving accuracy and precision for estimating the prevalence of rarer diseases. This is demonstrated by Figure 3, a mixture size of 500 gives imprecise estimates around a 10% disease prevalence (proportion 0.1) and includes zero. Increasing the mixture size to 5000 significantly improves the precision around the same 10% prevalence allowing a meaningful estimate of disease prevalence.

### What is the impact of the size of the mixture cohort?

With all but the smallest cohort sizes prevalence estimates remain valid. Not surprisingly, increasing cohort size leads to an improvement in the precision of estimates, (Fig. 3 and Fig. 4). Increasing mixture size improves precision because larger cohort sizes can be seen to represent the characteristics of the reference distributions more accurately. Where larger mixtures cohort sizes are not possible Figure 4 clearly demonstrates that for all methods except the Excess, accurate albeit more imprecise, estimates of disease prevalence can still be achieved with lower case numbers. Figure 3 shows that using a T1DGRS and a mixture of just 500 cases can still provide accurate and clinically informative estimates around a disease prevalence of 40%, e.g., determining prevalence of T1D in diabetes cases rapidly requiring insulin (clinical PPV of ≈50% for identifying T1D [12]).

### Can less discriminate genetic risk scores be used?

Accuracy and precision of estimates for all four methods reduces when using less discriminative GRS. However, excluding the excess method robust estimates of proportion are possible even when using GRS with AUC around 0.6. This is demonstrated in Figure 5 where we create artificial genetic risk scores with the area under the ROC curve (AUC) varying from completely non-discriminative (AUC = 0.5) to fully discriminative (AUC = 1). Reducing GRS AUC leads to widening of confidence intervals around disease prevalence’s of 10% and 25%. The reduction in precision can be entirely mitigated by increasing the mixture cohort size. This is emphasised by Table 1, which shows the minimum mixture size required to give an estimated precision of 0.1 (*CI*_*U*_ − *CI*_*L*_) around a prevalence of 0.1 with increasing AUC. For instance using the EMD method a mixture size of 25,500 (2,550 cases and 22,950 non cases) and an AUC of 0.6 allows robust precision around a 10% disease prevalence. It is worth noting that these examples use normal distributions with equal standard deviations, representing the majority of polygeneic GRS. The performance of the EMD and KDE methods, which are non-parametric, will be enhanced relative to the Means method when the reference cohort means are close but there are differences in other characteristics of the GRS distributions (e.g., skewness, kurtosis or multi-modality). This is the case, for example, when certain genetic variants predominate leading to a skewed GRS distribution e.g., in autoimmune diseases such as T1D with strong HLA association.

### What is the relative performance of the different methods?

We find that the Means, KDE and EMD methods perform well in estimating prevalence. Their accuracy and precision are largely comparable and all outperform the Excess method. The Excess method demonstrates reduced performance and exhibits strong bias (difference between the estimated prevalence 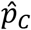 and the median of the bootstrap values 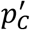) typically underestimating the true prevalence. Figure 5 shows that regardless of the mixture size, the Excess method is practically unusable for any but the highest AUC. The relatively comparable performance of the Excess method in Fig. 3 is a consequence of the high AUC of the T1DGRS (0.88) and the strong asymmetry of the reference distributions.

### Clinical example estimating prevalence of coeliac disease

Finally, we illustrate a worked example asking the question of how much undiagnosed coeliac disease is present within a population adhering to a gluten-free diet (Fig. 6) using a coeliac disease GRS (CDGRS). This is important as whilst people observe a gluten-free diet for a number of reasons, it is possible that without getting a formal diagnosis people with undiagnosed coeliac disease eliminated gluten from their diet using trial and error to alleviate abdominal symptoms. For each method we: 1) compute an estimate of prevalence 2) use modelled mixtures and bootstrapping to calculate its confidence intervals. All methodologies provide estimates of the proportion of individuals with coeliac disease with their 95% CIs shown in square brackets: Excess = 15.0% [12.1%, 16.7%]; Means = 15.1% [13.5%, 16.8%]; EMD = 15.1% [13.5%, 16.8%]; KDE = 13.2% [11.5%, 15.0%]. The 13.9% prevalence of the coeliac disease was calculated from reported cases in the UK biobank. Our results suggest an absence of undiagnosed coeliac disease in all patients adhering to a gluten-free diet and not known to have the condition.

**Figure 6:**
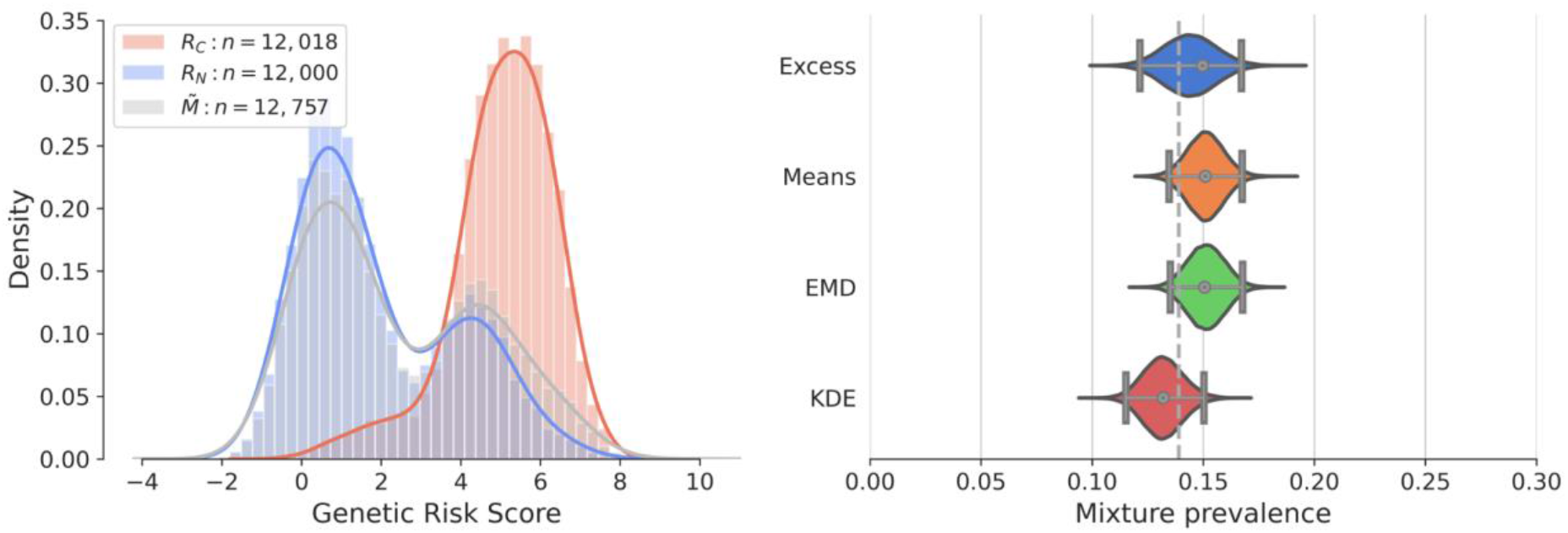
Coeliac disease dataset worked example. A comparison of the four methods applied to a gluten-free cohort from the UK biobank (mixture population 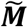). The reference and the mixture distributions are plotted on the left (*R*_*C*_, shaded red, *R*_*N*_, shaded blue, 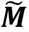, shaded grey, respectively). Estimated values of prevalence 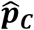 and 95% confidence intervals (grey dots and lines with vertical bars at the ends) are plotted on the right showing estimates of Excess = 15.0%, Means = 15.1%, EMD = 15.1%, KDE = 13.2%. The violin plots show the distribution of the 100,000 estimates of prevalence 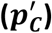 in the bootstrap cohorts. The proportion of participants adhering to a gluten-free diet and reporting coeliac disease is shown as a dashed vertical line. Sample sizes: non-cases UK Biobank (*n* = 12000), cases coeliac disease reference cohort (*n* = 12018), mixture self-reported gluten-free diet UK Biobank (*n* = 127*5*7).

## Discussion

We present analysis of a novel approach to disease classification based on genetic predisposition. We demonstrate genetic stratification produces robust prevalence estimates even in the context of: rarer diseases, smaller cohort sizes and less discriminative genetic risk scores, highlighting the general utility of the proposed approaches. This was demonstrated through head-to-head evaluation of four methods including the original Excess methodology published by Thomas et al. [7]. The presented examples illustrate the performance and utility of these method across a range of different scenarios highlighting improved accuracy of the new approaches over the original Excess method. We supplemented the estimation methods by combining Monte Carlo sampling [13] and bootstrap [14] methods to quantify uncertainty around the estimate and compute realistic confidence intervals.

### Distribution of genetic risk scores can be used to estimate population prevalence

Our results show that robust estimates of prevalence are possible using differences in distributions of genetic risk scores between a population of cases and non-cases. Our methods build on the previously published genetic stratification by Thomas et al. [7]. This novel concept is important, as when coupled to the ever-increasing availability of large population level genetic datasets, it allows fresh insights into disease epidemiology without requiring extensive investigations or unreliable self-reported diagnosis [5, 6]. The permanence associated with genetic risk makes these methods potentially very powerful tools for clinical researchers and enables accurate evaluation where cases are difficult to differentiate clinically.

### Large mixture cohorts are not required but improve precision of estimates

Accurate estimates were possible with mixture containing as few as 500 individuals with precision around estimates improved by increasing cohort size. The explanation for this is that sufficient numbers of individuals are required for the mixture to robustly represent the true distributions of cases and non-cases. Increasing the mixture cohort size can mitigate for the reduction in precision observed with decreased GRS discrimination (lower AUC). For instance a mixture size of 25,500 allows precision around a prevalence of 0.1 using a polygenic GRS with AUC 0.6. This represents 2,550 cases and 22,950 non cases, achievable for many polygenic disease in modern day population dataset, for example UK Biobank has genotyped ≈ 500,000 individuals [9].

### Rarer disease can be detected, with accuracy improved by cohort enrichment

Away from extremes of proportion the prevalence of the disease had little impact. The extent to which proportions were within the extremes reduced with increasing AUC of the GRS used. Moving the mixture composition away from the extremes can be achieved through mixture enrichment. For example, to increase the proportion of T1D within a cohort with diabetes, the mixture cohort could be restricted to include only insulin treated diabetes [7]. Mixture enrichment will inevitably be to the detriment of mixture size and therefore cause some reduction in precision around estimates, but this will be outweighed by improved accuracy.

### Estimates remain robust in diseases with less discriminative genetic risk scores

Whilst accuracy and precision are higher when utilising a more discriminative GRS, we show that clinically meaningful estimates can still be obtained using GRS with AUC as low as 0.6. Performance of the Means, EMD and KDE methods is very good in the case of normal GRS distributions with equal standard deviations e.g., diseases with polygenetic risk arising from a large number of causal variants, each with tiny effects e.g., T2D. In diseases where certain variants predominate e.g., HLA in autoimmune disease, the GRS will be skewed to account for this e.g., T1D. In this instance the EMD and KDE methods will be more accurate, as they are able to utilise the unequal skewness (or other properties such as standard deviations or kurtosis) even when the means of the reference distributions are close, see Supplementary Fig. 5. In diseases where one variant has the predominant effect on genetic risk e.g., HLA-DQ in coeliac disease, it would be possible to estimate prevalence using just this variant. However previous work has shown a GRS including the predominant as well as smaller effect variants has better discriminative ability than the predominant variant alone [15].

### Different methods have different advantages

In most settings the best approaches are the Means, EMD and KDE methods. The overall performance of these three methods is comparable across different parameters (mixture size, mixture proportional makeup and GRS AUC). At extreme proportions the KDE method exhibits the smallest bias. A key advantage of the Means method is that it is very straightforward to apply, allowing rapid evaluation of disease prevalence within a cohort. Alternatively, the EMD and KDE methods have the benefit of being able to estimate the prevalence in cases where the Means method cannot be used, e.g. if the reference cohorts have very similar means (Supplementary Fig. 5). Finally, the KDE and EMD methods can be used to test the assumption that the mixture is only composed of two cohorts (Supplementary Fig. 8).

As noted in the original article by Thomas et al. [7] the Excess method inherently underestimates the proportion of cases because typically both reference cohorts have values below the median value of *R*_*N*_. Taking distinct approaches, the new methods eliminate this inaccuracy and even with decreasing genetic discrimination these are still interpretable, reflecting the improved generalizability of these methods. We note that the Excess method could be modified to improve its accuracy (e.g., by choosing another quantile rather than the median) but these changes would require case-by-case fine-tuning and at best achieve equivalence to the proposed alternative methods.

### Utility of using polygenic approaches to estimate prevalence within a group

#### Prevalence

We highlight the clinical utility of the presented concept with a clinical question around coeliac disease. Exclusion of gluten from the diet is a treatment for coeliac disease. Coeliac disease can present with non-specific abdominal symptoms and diagnosis is often difficult [16]. We show that it is possible to determine if there is any undiagnosed coeliac disease within a cohort adhering to a gluten-free diet using our methods. This question would be unanswerable using the traditional clinical approach of endoscopy to confirm coeliac disease, as once observing a gluten-free diet findings are often normal [16]. We estimated the proportion of those reporting a gluten-free diet within biobank using the coeliac disease GRS and then compared this against all those reporting a diagnosis of coeliac disease in the same gluten-free cohort. The genetic estimates and those with a reported diagnosis of coeliac disease were comparable, suggesting that there is no undiagnosed coeliac disease within this gluten-free cohort. Whilst this finding is not unexpected, it could not be robustly shown before and highlights the general applicability of the proposed framework to quantitatively answer novel and difficult to answer questions.

#### Defining clinical characteristics of a genetically defined subgroup

A further advantage of the proposed methodologies over traditional clinical classification arises from the fact that clinical characteristics are not used to define cases. It is therefore possible to estimate both binary and continuous clinical characteristics of the genetically defined disease group within the mixture cohort. Using BMI as an example:

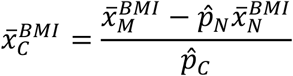

Where 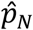 and 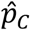 represent the estimated proportions and 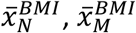 and 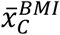 represent the mean BMI of each of the non-cases, mixture and cases (disease) groups respectively. This approach was used in [7] to show rates of Diabetic Ketoacidosis to be the same in T1D diagnosed above and below 30 years of age. We note that all the same limitations of the Means method apply. The EMD and KDE methods could allow for reconstruction of the full distribution of the clinical characteristic, however evaluation of this approach is beyond the scope of this study.

### Testing of proposed clinical discriminators

Another utility of these genetic discrimination techniques is to test the performance of clinical classification criteria and allow more precise stratification of a population. Whilst the increasing availability of population datasets generated from routinely collected data allows large scale population analysis, robust classification can become more difficult leading to bias which is difficult to quantify [6]. Treating the clinically defined cohort as a mixture would allow rapid estimation of the correctly and incorrectly classified proportions within the cohort, thus allowing for bias adjustment and optimisation of classification criteria.

### Cautions

The use of genetic data in the context of genetic stratification means certain assumptions must hold true for the estimates to be valid. The same assumptions required for Mendelian randomisation [1, 17] should be met here. Key to the accuracy of estimates is the equivalence assumption which states that cases and non-cases in the mixture reflect the respective reference cohorts. This is particularly important when studying different geographical populations where allele frequencies may vary, leading to an alteration in genetic risk scores across the cohorts. The assumption will also fail where the GRS is different between cases or non-cases in the reference and mixture. This is shown by our example of a higher T2DGRS distribution for a WTCCCC reference T2D population compared to T2D cases in UK Biobank, Supplementary Fig. 1. This reflected the WTCCC cohort being enriched for T2D risk variants as selected based on a positive family history of T2D or early disease onset [8]. This highlights the importance of testing the equivalence assumption prior to analysis. This should initially involve detailed assessment of the selection criteria for the mixture and reference cohort and available literature. Furthermore we suggest comparing the GRS of the reference non-cases (controls) and cases with, where available, the GRS of definite non-cases and cases within the same population the mixture has been taken from (e.g., by means of a statistical test appropriate for assessment of the observed GRS distributions).

Finally, all our methodologies assume that the mixture consists of only the two genetic reference cohorts such that *p*_*C*_ + *p*_*N*_ = 1. Both, the EMD and KDE methods provide a way to check if this mixture assumption is satisfied. In the case of the EMD method we could use the independent estimates of 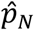 and 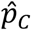 to check how much their sum deviates from 1. For the KDE method, the validation could be based on the residuals of the least-square fitting procedure. To check if the deviation from *p*_*C*_ + *p*_*N*_ = 1 is significant we again suggest the use of bootstrap methodology. We present some details and an example of such checks in the supplementary information, however detailed analysis of this aspect of the proposed methodology is beyond the scope of this paper.

## Conclusion

We propose novel approaches that use population distributions of genetic risk scores to estimate disease prevalence. We show that the proposed Means, EMD and KDE approaches improve upon the existing Excess method, performing similarly across different mixture cohorts, with robust estimates possible even when using GRS with reduced discriminative ability. Utilising these concepts will allow researchers to gain novel unbiased insights into polygenic disease prevalence and clinical characteristics, unhampered by clinical classification criteria.

## Online methods

### Participants

#### Type 1 diabetes cases

Cases (n=1,963) were taken from the Wellcome Trust Case Control Consortium [8]. The WTCCC T1D patients all received a clinical diagnosis of T1D at <17 years of age and were treated with insulin from the time of diagnosis.

#### Type 2 diabetes cases

Cases (n=1,924) were taken from the Wellcome Trust Case Control Consortium [8]. The WTCCC T2D patients all received a clinical diagnosis of T2D.

#### Coeliac disease reference cases

Cases (n=12,018) Cases consisted of those from a combination of European studies. Cases were diagnosed as previously described [18].

#### Coeliac non-cases

Non-cases (n=12,000) a cohort was randomly selected from those within the UK biobank (total n= 366,326) defined as unrelated individuals of white European descent without a diagnosis of coeliac disease and not reporting a gluten-free diet.

#### Gluten-free diet

Gluten-free cases (n=12,757) were taken from unrelated individuals of white European descent in the UK biobank reporting adherence to a gluten-free diet.

#### Reported coeliac cases in biobank

Coeliac disease cases (n=1,772) were defined based on self-reported questionnaire answers and/or an ICD10 record from hospital episode statistics data.

### Calculating genetic risk scores

#### T1DGRS

The T1DGRS was generated using published variants known to be associated with risk of T1D. All variants were present in the UK Biobank imputed genotype data. We followed the method as previously described [2] using tag variants rs2187668 and rs7454108 to determine HLA DR haplotype and ascertain the HLA-haplotype component of each individual’s score [19]. This was added to the score of the remaining variants, generated by summing the effective allele dosage of each variant multiplied by the natural log (ln) of the odds ratio.

#### T2DGRS

The T2DGRS was generated using published variants known to be associated with risk of T2D [20]. We generated a 77 SNP T2D-GRS in both the WTCCC cohort and UK Biobank consisting of variants present in both data sets and with high imputation quality (R2>0.4). The score was generated by summing the effective allele dosage of each variant multiplied by the natural log (ln) of the odds ratio.

#### CDGRS

The 46 SNP coeliac GRS was generated using published variants known to be associated with risk of Coeliac disease [21], [18], [22]. The log-additive CDGRS was generated using a weight as the natural log of corresponding odds ratios. For each included genotype at the DQ locus, the odds ratio was derived from a previously described case-control dataset [18]. For each non-HLA locus, odds ratios from existing literature were used, and each weight was multiplied by individual risk allele dosage.

### Excess method

Following on from previous work [7], the Excess method calculates the reference proportions in a mixture cohort according to the difference in expected numbers either side of the reference cohort’s median. The reference median in question was taken to be the closest to the mixture cohort’s median. The proportion was then calculated according to: 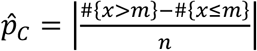, where *m* is the median of the reference cohort, *n* is the size of the mixture cohort and *x* is an individual participant in the mixture cohort, hence #{*x* > *m*} represents the number of cases above the median and #{*x* ≤ *m*} represents the number of cases below the median.

### Means method

The mean genetic risk scores were computed for each of the two reference cohorts and the mixture population. The proportions of the two reference cohorts were then calculated according to the normalised difference of the mixture cohorts’s mean 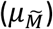 and the means of the two reference cohorts (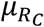 and 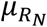): 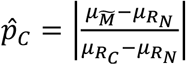. If the mean of the mixture cohort is bigger (or smaller) than both means of the reference cohorts then the estimate is defined as 1 (or 0) depending on the closest reference mean.

### Earth Mover’s Distance (EMD) method

Intuitively, the Earth Mover’s Distance (EMD) is the minimal cost of work required to transform one ‘pile of earth’ into another; with each ‘pile of earth’ representing a probability distribution. Mathematically, the (EMD) is a Wasserstein distance and has been widely used in computer and data sciences [23, 24]. For univariate probability distributions, the EMD has the following closed form formula [25]:

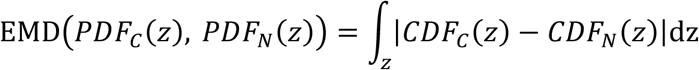

Here, *PDF*_*C*_ and *PDF*_*N*_ are two probability density functions with support in set *Z*, and cumulative density functions, *CDF*_*C*_ and *CDF*_*N*_, are their respective cumulative distribution functions.

To compute the EMD, we first find the experimental CDFs of genetic risk scores for each of the two reference cohorts and the mixture cohort. These CDFs are then interpolated at the same points for each distribution, with the points being the centres of the bins obtained when applying the Freedman-Diaconis rule [26] to the combined reference cohorts (such that 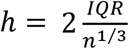. As a support set, we take an interval bounded by the minimum and maximum value of the genetic risk score in all three cohorts. The proportions were then calculated as: 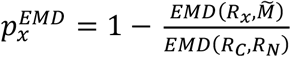, where *x* is either *c* or *N*. Since the two estimates are independent, deviation of their sum from one, 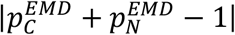 can be used to test the assumption that *p*_*C*_ + *p*_*N*_ = 1, dispersion of the deviation can be computed during bootstrapping and compared with the value observed in the analysed cohort. However, under the assumption that *p*_*C*_ + *p*_*N*_ = 1, we adapted the method by taking the average of the estimated proportions as follows: 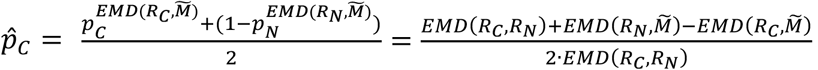.

### Kernel Density Estimation (KDE) method

Individual genetic risk scores were convolved with Gaussian kernels, with the bandwidth set to the bin size obtained when applying the Freedman-Diaconis rule [26] in the same way as for the EMD method. This forms two reference distribution templates and a mixture template, *KDE*_*C*_, *KDE*_*N*_ and *KDE*_*M*_ for each dataset. A mixture model was then defined as the weighted sum of the two reference templates (with both weights initialised to 1). This model was then fit to the mixture template (*KDE*_*M*_) with the Levenberg-Marquardt (Least Squares) algorithm [27], allowing the weights (*w*_*C*_ and *w*_*N*_) to vary. The proportions were then calculated according to: 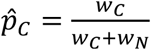. Admissible values of the weights are limited to the [0,1] interval.

### Simulated mixtures

To simulate a range of real-world scenarios, we constructed artificial distributions by randomly sampling (with replacement) genetic risk scores from the reference cohorts of cases (*R*_*C*_) and non-cases (*R*_*N*_) in specified proportions of each reference cohort, *p*_*C*_ and total mixture sizes, *n*. To simulate mixtures, we use the Wellcome Trust Case Control Consortium [8] T1D (n=1,963) and T2D (n=1,924) data. We used half of the available samples as reference cohorts (first n=982 and n=964 points, respectively) and the other half (last n=981 and n=964, respectively) is a hold-out set used to construct the mixtures. To obtain any required mixture size we sampled with replacement from the hold-out data.

For the heatmaps, Fig. 4 and Supplementary Figs 2–4, the proportion and cohort size were systematically varied, with *p*_*C*_ ranging from 0 to 1 in 0.01 (1%) steps while *n* ranged from 100 to 2,500 in steps of 100 samples. All four methods were applied to each combination of these parameters. At each point in the parameter space, we estimated the prevalence 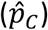 and its confidence interval and then compared it with the model proportion (*p*_*C*_) used to generate them.

Figure 4 (top row) illustrates how the randomness of the simulated mixture cohort affects the variability of each method’s estimates. This variability reflects the randomness that is inherently present in the mixture cohort. Supplementary Fig. 2 shows how this variability decreases for more discriminative GRS, while Supplementary Figs 3 and 4 compare the performance of the methods once the randomness of the composition of the mixture cohort is eliminated.

### Synthetic GRS data

To generate synthetic GRS in Fig. 5 and Supplementary Fig. 5 we used pseudorandom number generators. As references, we used two samples (n=2,000, each) from normal distributions with mean 0 and standard deviation 1, *N(0,1)*; the means and standard deviations of the reference samples were µ = −0.002, σ = 0.999 and μ = −0.008, σ = 1.001. The reference samples are generated only once. To change the AUC for the reference samples, we added a value to one distribution of them to change its mean. The mixtures are generated using different pseudorandom number generators for each proportion (*p*_*C*_) and AUC value. For example, to generate mixture with n=5,000, *p*_*C*_ = 0.1 and AUC = 0.7 we: 1) draw 500samples from *N(0,1)* and 2) we draw n=4,500 samples from *N(0,1)* and add 0.74 to them. The mixture and reference samples are generated separately.

### Varying mixture size

To investigate dependence of the width of the CIs on the mixture size (Fig. 5C) and find the minimum mixture size required for CIs width <0.1 (Table 1) we used the same synthetic GRS distributions with *p*_*C*_ = 0.1 as described above. For the Excess, Means and the EMD methods, we varied mixtures sizes between 100 and 10,000 (30,000 for AUC <0.7) with a step of 100 points. Since the KDE method is more computationally expensive, we tested mixture sizes between 100 and 6,500 with a step of 100 points and between 7,000 and 10,000 (40,000 for AUC=0.6, 30,000 for AUC=0.65) with a step of 500 points. For each considered mixture size, we repeated estimation of the CIs 100 times. We disregarded estimates for which the CIs do not include *p*_*C*_ = 0.1. As the minimum mixture size, we took a median of the mixture sizes (over the 100 runs) at which we first observed the CI width <0.1.

### Calculating confidence intervals

In order to estimate confidence intervals and any systematic bias of the methods we used Monte Carlo [13] and bootstrap methods [14, 28]. We combined the two approaches in order to capture variability of the estimate resulting from the mixture size and features of the reference distributions.

First, we stochastically modelled the process of generating the mixture. To do so, we modelled *N*_*M*_ new mixtures, by sampling with replacement from the reference cohorts. Each modelled mixture has the same size as the original cohort and the composition given by the initial estimate 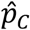 based on the original mixture. For example, if the original cohort has 1,000 values and the estimate was 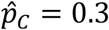 then each modelled mixture would contain 300 values sampled with replacements from the cases reference sample (*R*_*C*_) and 700 values from the non-cases reference sample (*R*_*N*_). Next, we resampled each of the *N*_*M*_ new mixtures generating *N*_*B*_ bootstrap samples, see also Supplementary Fig. 7.

Following, chapters 2 and 5 from [14] we used all the *N*_*M*_ · *N*_*B*_ cohorts to compute the bias and confidence intervals of the estimate. The systematic median bias of the method is defined as a difference between med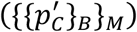 the median value of the *N*_*M*_ · *N*_*B*_ bootstrapped estimates of 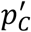 and the estimate 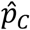:

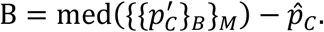

We used bias corrected and accelerated bootstrap confidence intervals (BCa CI) which we computed as described in [29].Bootstrap confidence intervals are based on the assumption that the spread of the distribution of the bootstrap estimates 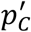 can be used to estimate the CI. The BCa CI take into account median bias and skewness (acceleration) of the distribution of the bootstrap estimates 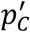 and allows calculation of corrected quantiles representing a chosen confidence level, α.

Throughout this section Φ is a normal standard (µ = 0, σ = 1) CDF and Φ^−1^ is its inverse and 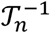 is an inverse CDF of a Student’s *t*-distribution with *n* degrees of freedom. The computation takes the following steps:

1. Estimate the median bias correction factor *z*_0_:

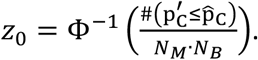
2. Estimate the acceleration correction factor *â*:

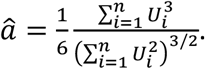 Where *U*_*i*_ values are calculated using the *jackknife influence function*:

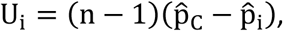 Here 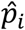 is an estimate based on the reduced mixture 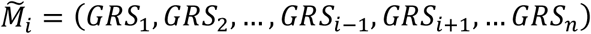 with score *i* removed.
3. To counteract the narrowness bias we additionally expand the confidence level [28]

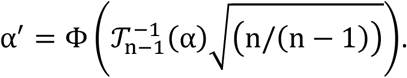
4. Use bias and acceleration factors to compute the BCa confidence levels:

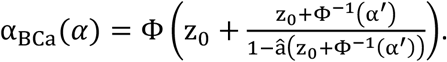
5. Take *α*_*Bca*_(*α*/2) quantile of the 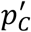 samples to obtain the lower confidence limit *CI*_*L*_ and *α*_*Bca*_(1 − *α*/2) quantile to obtain the upper confidence limit *CI*_*U*_. If the median bias is very strong the BCa CI are undefined. For example, if the 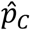 is outside of the range of the distribution of the bootstrap estimates 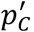, *z*_0_ is infinite and both limits of the *CIs* are equal to the maximum or minimum value the 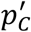 samples.

## Data Availability

The software implementing these methods is open-source and available under version-control here: https://github.com/bdevans/DPE.
UK Biobank data is a open access resource

## Acknowledgements

This research has in part been conducted using the UK Biobank Resource.

The authors would like to acknowledge the use of the University of Exeter High-Performance Computing (HPC) facility in carrying out this work.

We are grateful to Jack Bowden for his comments on the manuscript.

## Author contributions

Manuscript writing: BDE, NJT, PS, KTA

Method development and implementation: BDE, PS, NJT, ATH, RJO, KTA

Data acquisition and coding: NJT, SS, RK, SJ

Simulation running and analysis: BDE, PS

Discussion of results and manuscript editing: All authors Project coordination: KTA, NJT

## Footnotes

1 The software implementing these methods (in Python and Matlab) will be open-sourced upon acceptance through a public version-controlled code repository and linked to here.

