## Supplementary material for "Estimating population level disease prevalence using genetic risk scores": supplmentary

**Supplementary information**

**Supplementary Text and Figures**

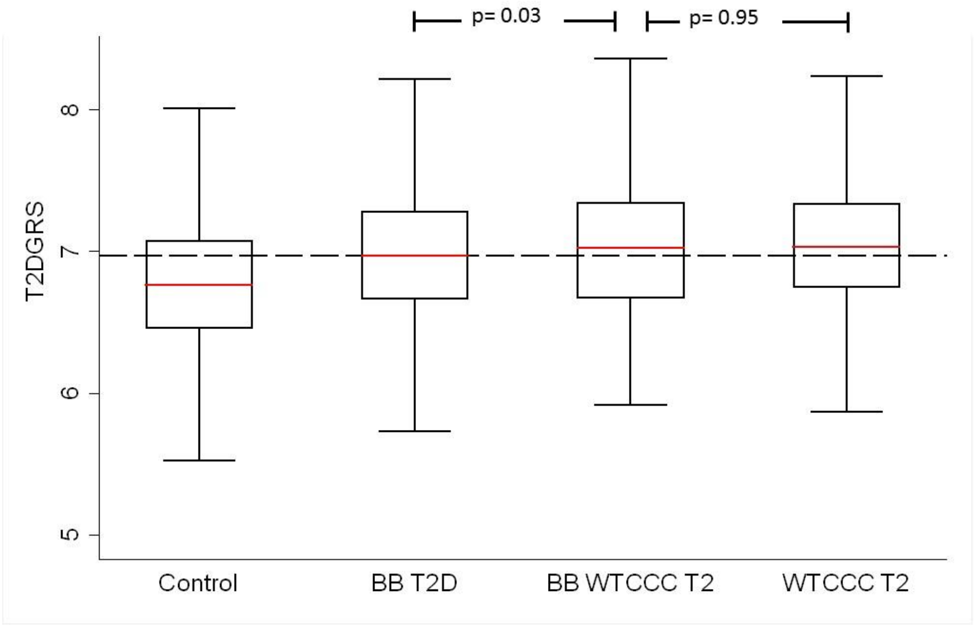


**Supplementary Figure 1: shows the effect of non-preserved equivalence. BBT2D = Biobank Type 2 diabetes defined as diabetes diagnosed over 30 years of age and non-insulin treated. BB WTCCC T2 is defined as first degree relative with type 1 diabetes diagnosed aged over 30 up to 40 years of age to recreate the WTCCC cohort. The WTCCC T2 cohort is also shown.**

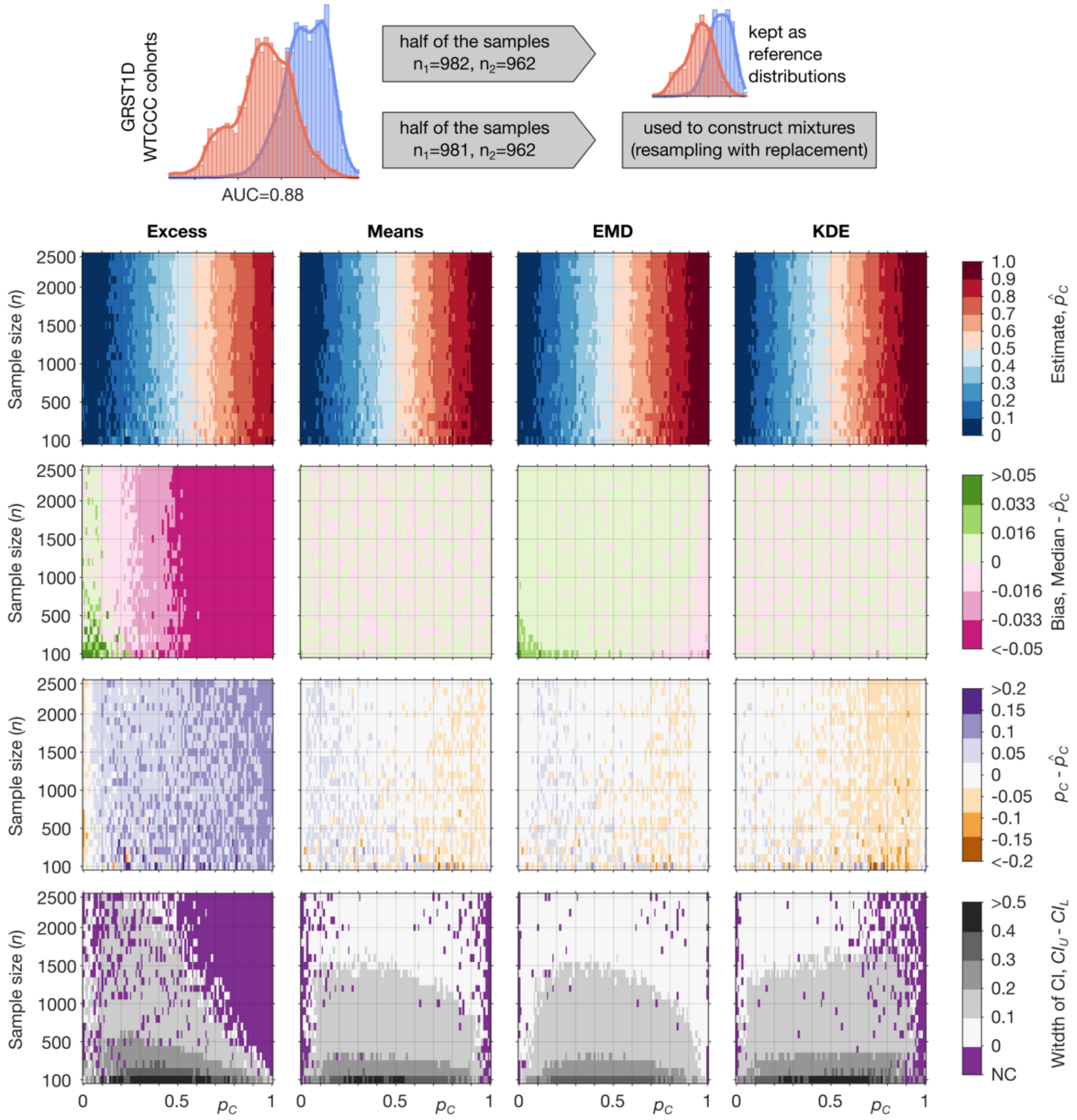


**Supplementary Figure 2: A comparison of the four methods prevalence estimates and confidence intervals for varying proportion of disease and cohort sizes using the T1DGRS from the WTCCC dataset ($n = 3887$). (Top row) Estimate of prevalence ($\hat{p}_C$) in the constructed mixtures. (Second row) Bias of the prevalence estimates ($\hat{p}_C$) across the constructed mixtures. (Third row) deviation from the true proportion ($p_C - \hat{p}_C$) across the constructed mixtures. (Bottom row) The width of confidence ($CI_U - CI_L$) intervals of the estimates across the constructed mixtures. The purple colour (bottom row) indicates regions in which the confidence interval did not include the true value ($p_C$), $CI_U = CI_L$ or CI are undefined (both latter cases can happen if $\hat{p}_C = 0$ or $\hat{p}_C = 1$). Sample sizes: $R_C$ – cases WTCCC type 1**

**diabetes ($n = 982$), $R_N$ – non-cases WTCCC Type 2 diabetes ($n = 962$), mixtures – sampled with replacement from a holdout half of the $R_C$ ($n = 981$) and $R_N$ ($n = 962$) samples.**

### Quantitative characterisation of methods

To compare bias and precision (quantified as the width of CI) of all the methods we compared their performance assuming that $p_C = \hat{p}_C$ and acceleration=0. Again, the proportion and sample size were systematically varied, with $p_C$ ranging from 0 to 1 in 0.01 (1%) steps while $n$ ranged from 100 to 2,500 in steps of 100 samples. All four methods were applied to each combination of these parameters. At each point in the parameter space, we estimated the bias and confidence intervals. This idealised scenario allows a direct head-to-head comparison of accuracy between all four methods without the randomness originating from the sampling process. Results of this comparison are presented in Supplementary Figs 3 and 4.

Supplementary Figs 3 and 4 show:

- (1st row) Median of the 100,000 bootstrap samples $\mathrm{med}(\{\{p'_C\}_{1000}\}_{100})$,
- ($2^{nd}$ row) Median bias $\mathrm{B} = \mathrm{med}(\{\{p'_C\}_{1000}\}_{100}) - \hat{p}_C.$,
- ($3^{rd}$ row) The width of confidence ($CI_U - CI_L$) intervals,

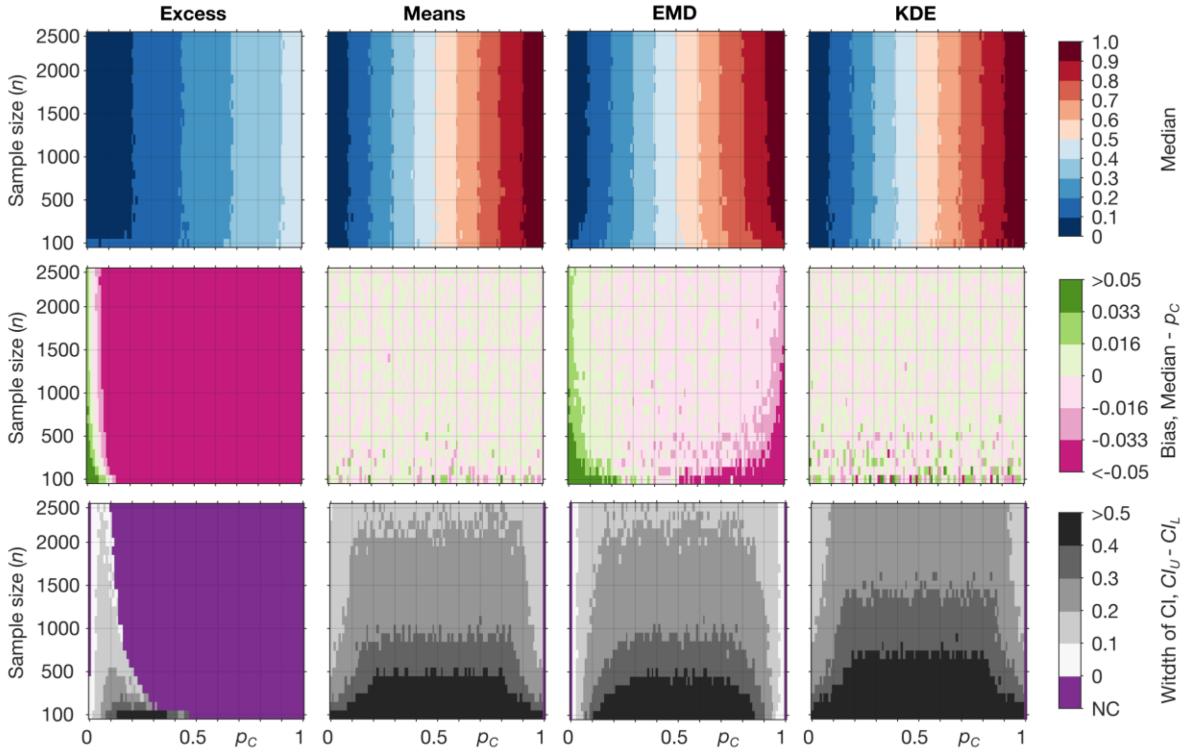


**Supplementary Figure 3: Evaluation of the four Estimation Methods using T2DGRS from the WTCCC dataset ($n = 3887$). (Top row) Median value of the 100,000 estimates of prevalence ($p'_C$) in the bootstrap samples across defined mixture proportions ($p_C$ ) and the mixture sample size ($n$) of the dataset for each method. (Second row) Median bias of the methods across the constructed samples. (Bottom row) The width of confidence intervals ($CI_U - CI_L$) of the individual estimates across the defined mixture proportions ($p_C$ ) and the mixture sample size ($n$). The purple colour (row 3) indicates regions in which the confidence interval did not include the true value (only observed for the Excess method), $CI_U = CI_L$ or CI are undefined (both latter cases can happen if $p_C = 0$ or $p_C = 1$). It can be observed that across the parameter space, the Means, EMD and KDE methods all typically outperform the Excess method. It is also evident that the Means and KDE methods practically do not exhibit any bias. A further increase of sample sizes would be recommended in order to reduce the width of the $CI$ below 10% (see Table 1). Sample sizes: $R_C$ – cases WTCCC type 1 diabetes ($n = 982$), $R_N$ – non-cases WTCCC Type 2 diabetes ($n = 962$), mixtures – sampled with replacement from a holdout half of the $R_C$ ($n = 981$) and $R_N$ ($n = 962$) samples.**

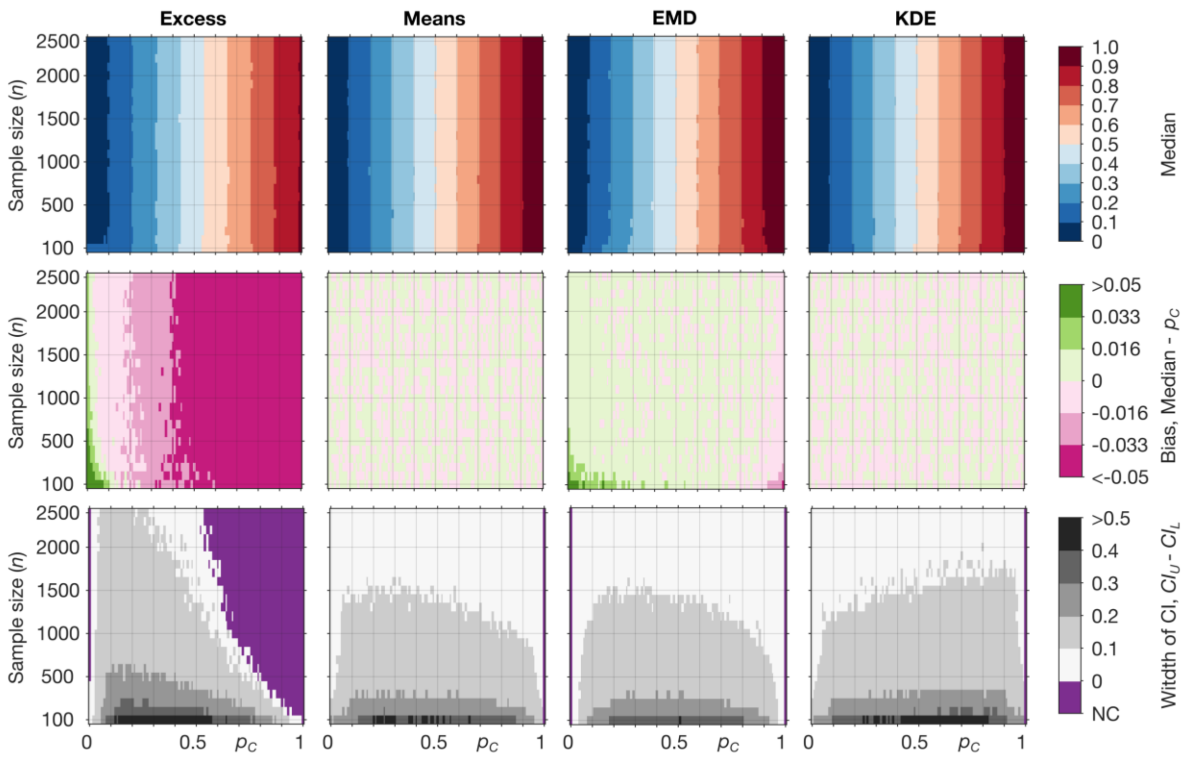


**Supplementary Figure 4: Evaluation of four Estimation Methods using the T1DGRS from the WTCCC dataset ($n = 3887$). (Top row) Median value of the 100,000 estimates of prevalence ($p'_C$) in the bootstrap samples across defined mixture proportions ($p_C$ ) and the mixture sample size ($n$) of the dataset for each method. (Second row) Median bias of the methods across the constructed samples. (Bottom row) The width of confidence intervals ($CI_U - CI_L$) of the individual estimates across the defined mixture proportions ($p_C$ ) and the mixture sample size ($n$). The purple colour (row 3) indicates regions in which the confidence interval did not include the true value (only observed for the Excess method), $CI_U = CI_L$ or CI are undefined (both latter cases can happen if $p_C = 0$ or $p_C = 1$). It can be observed that across the parameter space, the Means, EMD and KDE methods all typically outperform the Excess method. Sample sizes: $R_C$ – cases WTCCC type 1 diabetes ($n = 982$), $R_N$ – non-cases WTCCC Type 2 diabetes ($n = 962$), mixtures – sampled with replacement from a holdout half of the $R_C$ ($n = 981$) and $R_N$ ($n = 962$) samples.**

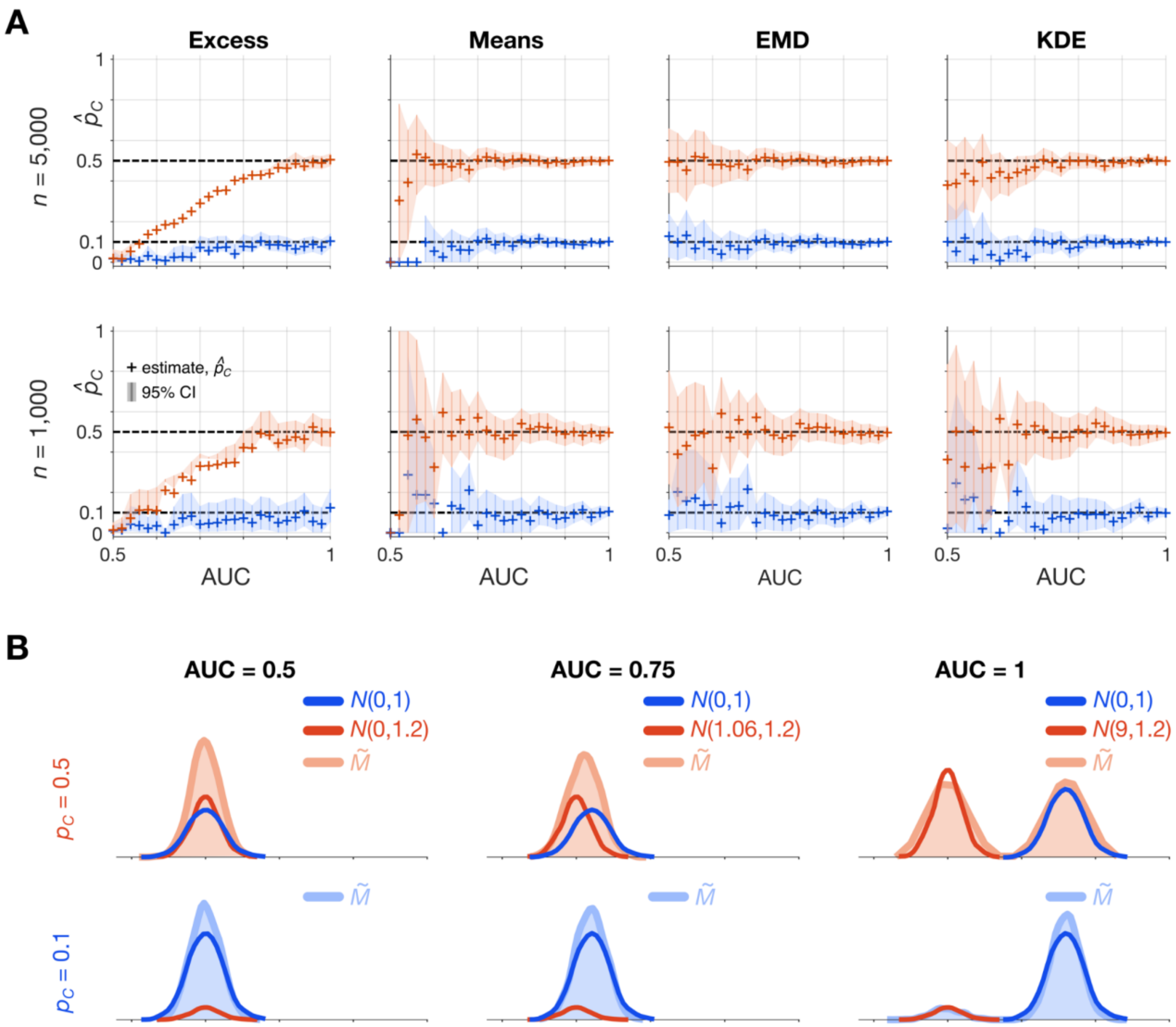


**Supplementary Figure 5: A comparison of the four methods using an artificial genetic risk score with increasing discriminative ability as measured by AUC, from AUC = 0.5 (no discriminative ability) through to AUC = 1 (complete differentiation). The estimated proportion with confidence intervals for each of the methods (Excess, Means, EMD, KDE) are shown using mixture sample size, $n = \{1000, 5000\}$. This figure is generated using artificial data: *N*(μ,σ) is a normal distribution with mean μ = {0.0, 0.093, 0.17, 0.25, 0.33, 0.41, 0.49, 0.57, 0.65, 0.4, 0.82, 0.91, 1.01, 1.1, 1.21, 1.31, 1.43, 1.55, 1.68, 1.83, 2, 2.17, 2.42, 2.73, 3.2, 9} and standard deviation σ={1, 1.2} and $\widetilde{M}$ is a mixture of the two normal distributions; $p_C = 0.1$ (blue) or $p_C = 0.5$ (red). Both reference samples have $n = 2000$. At several small AUC values, mean of the constructed mixture samples was smaller than both means of the reference samples, in these cases the prevalence estimate from the Means method is assumed to be $\hat{p}_C = 0$ and confidence intervals are undefined due to undetermined acceleration value.**

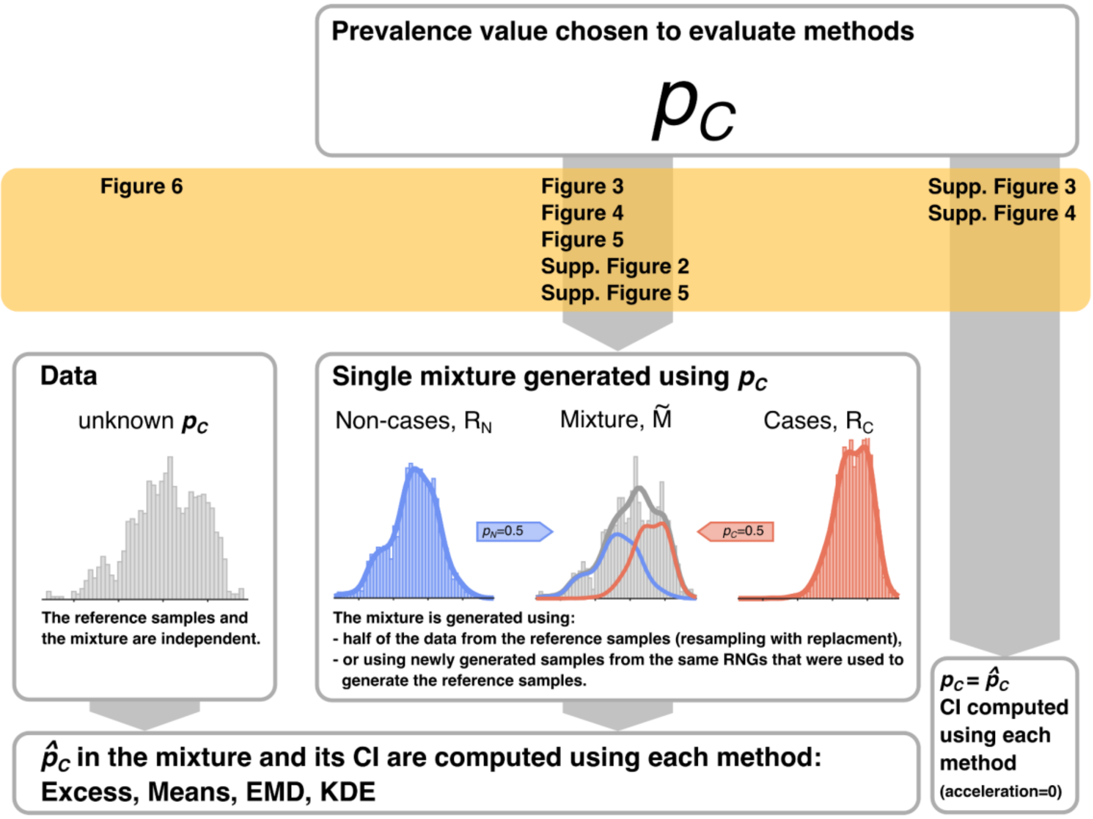


**Supplementary Figure 6: Illustration of the approaches used to estimate $\widehat{p}_C$ throughout the paper. The $\widehat{p}_C$ can be estimated from data (real or simulated) or can be fixed by hand. We used simulated data generated with a specified value of $p_C$ to evaluate the methods by comparing the estimated value, $\widehat{p}_C$, with the true prevalence, $p_C$ (results illustrated in Figs 3, 4 and 5, Supplementary Figs 2 and 5). In Supplementary Figs 3 and 4 with assume that $p_C = \widehat{p}_C$. In this way we compare bias and width of the *CI* of the methods without the random effects caused by simulating mixture data.**

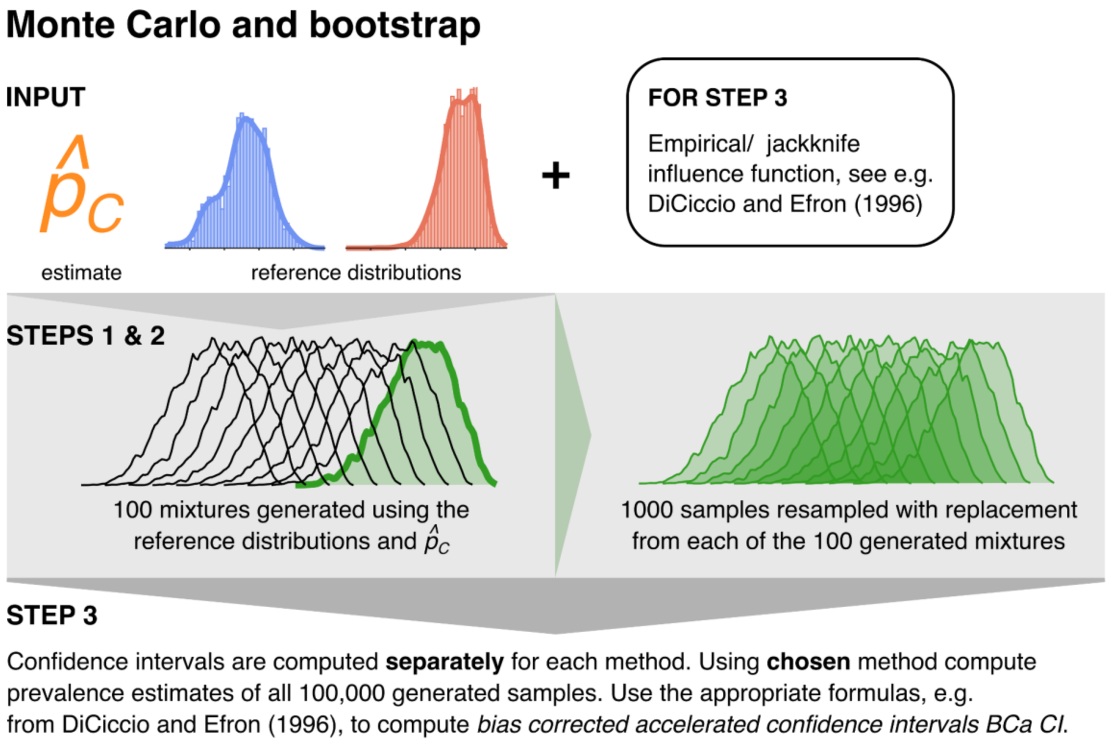


**Supplementary Figure 7: Illustration of the steps used in estimation of the confidence intervals of the prevalence estimate $\hat{p}_C$. To find bias corrected and accelerated (BCa) confidence intervals, we used the estimate $\hat{p}_C$, sample size of the original mixture sample, and the reference samples. We generated $N_M = 100$ mixture samples with a given composition ($\hat{p}_C$ ) and sample size ($n$) equal to the size of the original mixture sample. Next, we resampled (with replacement) each of the $N_M = 100$ new mixtures, generating $N_B = 1,000$ bootstrap samples for each. We applied each chosen method to all generated samples to obtain $N_M \cdot N_B = 100,000$ bootstrapped estimates $p'_C$. We then used the methods described in section "Calculating confidence intervals" to find BCa confidence intervals.**

### The mixture assumption

To check if the mixture assumption, $p_C + p_N = 1$, is satisfied, three different errors of the point estimates of prevalence derived from the EMD and KDE methods, $e_{EMD,1}$, $e_{EMD,2}$, $e_{KDE}$ could be further analysed. The $e_{EMD,1}$ error captures the deviation of the mixture from the convex combination of the two reference samples. The $e_{EMD,2}$ error is the EMD between the mixture and a model cumulative density function (CDF) based on the two independently estimated not normalised prevalence values $p_C^{EMD}$ and $p_N^{EMD}$. The $e_{KDE}$ error is the sum of squared residuals (multiplied by the Gaussian kernels bandwidth) from the least-squares fitting procedure, which forms part of the KDE method. In order to interpret the initial point values of the errors, we compared them to 100,000 bootstrapped error values (as in all other computations we used 100 mixtures * 1,000 bootstraps). The bootstrap samples are generated using the two reference samples and their composition is based on the estimate of prevalence. In this way, we compare the error value of the investigated mixture sample with 100,000 values from a model that *explicitly* assumes there are only two reference populations (i.e. $p_C + p_N = 1$). This approach allows us to check how likely the occurrence of the observed error value is in the model for a given sample size and reference samples. The obtained bootstrap *p*-values are the number of bootstrapped modelled errors that are higher than the sample error and can be interpreted as the probability that the observed values of $e_{EMD,1}$, $e_{EMD,2}$ and $e_{KDE}$ are a result of the sampling error. The bootstrap *p*-values are equivalent to *p*-values of a traditional statistical test (7, 11).

Supplementary Figure 8 shows an example of a mixture of two samples ($p_C + p_N = 1$) and an example of a mixture of three samples (with the third sample constituting 7.5% of the mixture, $p_C + p_N + 0.075 = 1$). It illustrates how the $e_{EMD,1}$, $e_{EMD,2}$ and $e_{KDE}$ errors could be used to test the assumption that the mixture is composed of only two samples. In the first case where the mixture is composed of just two samples, the observed $e_{EMD,1}$, $e_{EMD,2}$ and $e_{KDE}$ values are small, and when compared with the 100,000 bootstrapped error values, they indicated that there is a high chance: *p*=0.19 ($e_{EMD,1}$), *p*=0.95 ($e_{EMD,2}$) and *p*=0.96 ($e_{KDE}$) of observing them due to the sampling error in the mixture sample. In the second case where the mixture is composed of three samples, comparison of the observed $e_{EMD,1}$, $e_{EMD,2}$ and $e_{KDE}$ values with the bootstrapped values shows that they are unlikely to be a result of the sampling error: *p*=0.0001 ($e_{EMD,1}$), *p*<1e-5 ($e_{EMD,2}$) and *p*=0.003 ($e_{KDE}$). In fact, the value of $e_{EMD,2}$ is smaller than any of the bootstrapped error values. However, the figure shows only one particular example and the performance of the methods will depend on the mixture composition (contribution of the other sample) and features of the reference and the other samples.

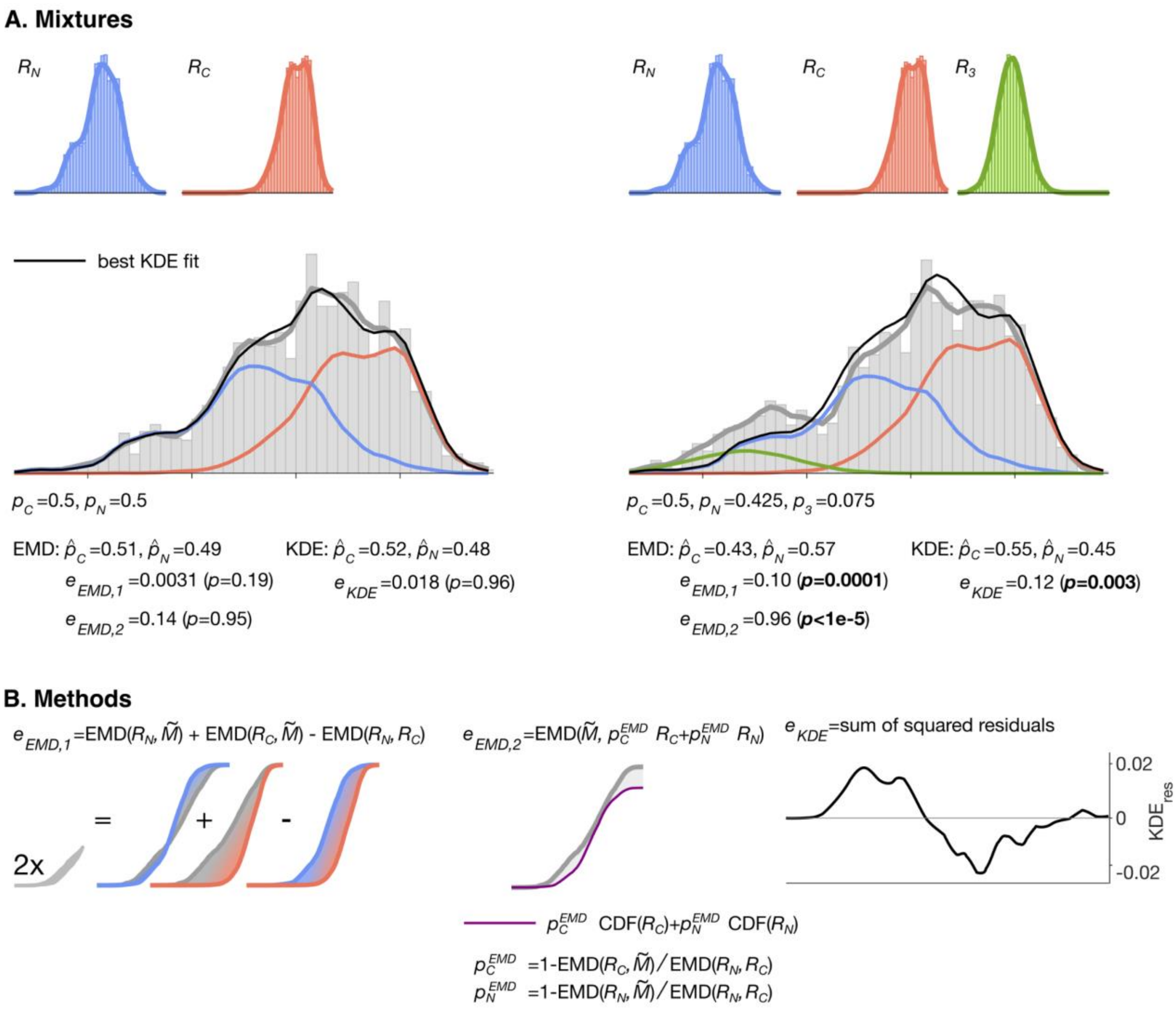


**Supplementary Figure 8: Worked examples of checking the mixture assumption, $p_C + p_N = 1$. A: An example of a mixture that consists of two samples and an example of a mixture that consists of three samples. Left: mixture of two reference samples ($p_C = 0.5$, $p_N = 0.5$). Right: mixture where the third sample has a small contribution ($p_C = 0.5$, $p_N = 0.425$, $p_3 = 0.075$; $R_3$ is a truncated normal distribution with mean 0.17 and std 0.025). B: Illustration of the methods: $e_{EMD,1}$, deviation from collinearity between the two reference distributions and the mixture; $e_{EMD,2}$, EMD between the mixture and a model CDF based on the two independently estimated prevalence values; $e_{KDE}$, the sum of squared residuals of the final fit.**
